# TG468: A Text Graph Convolutional Network for Predicting Clinical Response to Immune Checkpoint Inhibitor Therapy

**DOI:** 10.1101/2023.06.12.23291262

**Authors:** Kun Wang, Jiangshan Shi, Xiaochu Tong, Ning Qu, Xiangtai Kong, Shengkun Ni, Jing Xing, Xutong Li, Mingyue Zheng

## Abstract

Immunotherapy has achieved significant success in tumor treatment. However, due to disease heterogeneity, only a fraction of patients respond well to immune checkpoint inhibitor (ICI) treatment. To address this issue, we developed a Text Graph Convolutional Network (Text GCN) model called TG468 for clinical response prediction, which uses the patient’s whole exome sequencing (WES) data across different cohorts to capture the molecular profile and heterogeneity of tumors. TG468 can effectively distinguish survival time for patients who received ICI therapy and outperforms single gene biomarkers and TMB, indicating its strong predictive ability for the clinical response of ICI therapy. Moreover, the prediction results obtained from TG468 allow for the identification of immune status differences among specific patient types in the TCGA dataset. This rationalizes the model prediction results. Overall, TG468 could be a useful tool for predicting clinical outcomes and the prognosis of patients treated with immunotherapy. This could further promote the application of ICI therapy in the clinic.

## INTRODUCTION

Treating cancer more effectively remains a major challenge to human health. In recent years, immunotherapy has brought new hope to patients. However, due to the heterogeneity of tumors, the response to immune checkpoint inhibitor (ICI) drugs varies widely among different patients. While these drugs have achieved good effects in some patients, 60%-80% of patients do not have a clinical response to them[1].

The problem of determining a patient’s clinical response to ICI therapy could be alleviated by drug-related biomarkers. Recently, some predictive single gene biomarkers for ICI response have been reported, such as PAPPA2 and EPHA7 [2, 3]. The FDA has approved the use of the anti-Programmed Death Receptor 1 (PD-1) drug pembrolizumab in adults with high tumor mutational burden (TMB)—the number of somatic mutations per DNA megabase (Mb)— [TMB-H; ≥10 mutations/megabase (mut/Mb)] as a condition [4, 5], and the role of TMB as an independent biomarker of response to ICIs across different types of cancer is under investigation [4, 6-10]. However, currently reported clinical response biomarkers for ICIs have limited predictivity. Molecular characterization of frequent cancers has shown that these entities have high genetic complexity, and their clinical response is difficult to predict by single-gene biomarkers [11]. For example, some single-gene biomarkers that have been reported for pan-cancer prediction show poor performance in differentiating patients’ survival time, as evaluated by the *P* value of the log-rank test [3, 12, 13]. Furthermore, studies have demonstrated that TMB-H fails to predict ICI therapy response across all cancer types [7]. For example, Lyu et al. used a combination of multiple mutant genes to construct a linear model to predict TMB [14], while the model’s performance depends on the correlation of ICIs response with TMB, and the simple linear model cannot fit the data well. Therefore, more predictive biomarkers need to be developed to accurately stratify patients and identify those who will benefit most from ICI therapy[15].

Drawing inspiration from natural language processing (NLP) methods in artificial intelligence, we can treat the multiple genes of a patient as “words” and their combinations as “sentences”. These gene sentences can be combined to form a corpus of all patients in different cohorts. We can then use the patient’s clinical response to ICI drugs as the “label” of such a sentence. Just as in text classification, the drug response label corresponds to the classification label of a sentence. Therefore, we can use algorithms from the NLP field to explore the collective predictive performance of multiple gene biomarkers for ICI drugs, i.e., to formulate the drug response prediction problem into a text classification problem.

It should be noted that natural language has a grammatical structure and word order, making it difficult to determine how the patient’s gene “word” arrangement in a “sentence” may affect the semantic “label” of the sentence. To address this issue, we need to choose a method that ignores the effect of word order. One such model is the Text Graph Convolutional Network (GCN) [16], which can classify sentences regardless of word order. It constructs sentences and words as nodes in a heterogeneous graph, where the edges between word nodes and sentence nodes are determined using point-wise mutual information (PMI), and the edges between sentence nodes are determined using inverse document frequency (IDF) of the word in the document. Based on this model, we designed a model that treats the combination of genes as a sentence and predicts the probability of clinical response to ICI therapy. Since each patient’s gene sentence is constructed using a maximum of 468 genes, we named our model TG468, where TG stands for Text GCN.

In this way, TG468 transforms drug response prediction into a graph neural network-based text classification problem. The interaction between gene nodes allows the model to consider complex biological interactions, such as synergy or antagonism between genes’ status. Additionally, the overall graph composition of the model allows the model to consider the interaction between patient nodes and the interaction between cohorts of different tumor types. This feature forms the basis for learning the common molecular characteristics of immune checkpoint inhibitor (ICI) drug effects.

TG468 has achieved better clinical benefit prediction than single-gene biomarkers and TMB-H, that is, the log-rank test *P* values of differentiating patient survival time are more significant, and the hazard ratio (HR) coefficient is lower. Further analysis of the TG468 prediction results of The Cancer Genome Atlas (TCGA) data shows that TG468 can distinguish the difference in immune status between patients, demonstrating that prediction is reasonable. As whole genome sequencing (WES) remains an expensive and time-consuming method to capture the molecular profile and heterogeneity of tumors [5], our developed TG468 method has better clinical application value as it requires sequencing information of fewer genes and can balance the reliability and cost of ICI response prediction better.

## METHODS AND MATERIALS

### Model construction

First, we obtained the candidate gene set according to the process illustrated in Fig. 1a. We divided the patients in the reference set into responder (R) and non-responder (NR) groups. The response group includes patients with complete response (CR) and partial response (PR), while the non-response group includes patients with stable disease (SD) and progressive disease (PD). Gene mutation is defined as nonsynonymous somatic mutation. For each gene in the WES gene set of the reference set, Fisher’s exact test was performed to explore the relationship between gene status (mutated or wild type) and clinical benefit (R and NR) in response to ICI therapy. A candidate gene set of 983 genes with *P* values less than 0.05 was identified as potentially characterizing the patient’s clinical response.

**Fig. 1.**
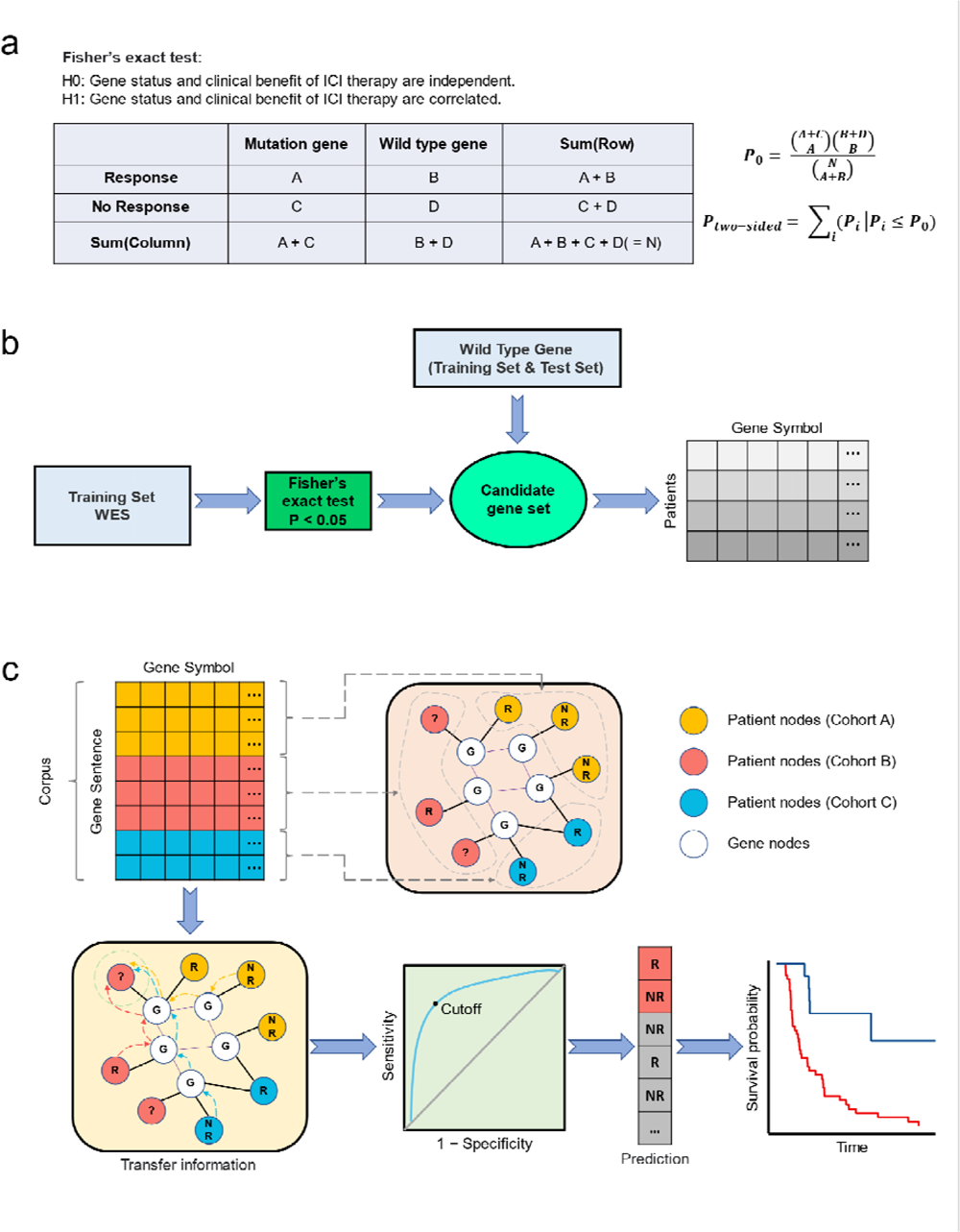
Flow chat of the study. **a,** Identify candidate genes. Fisher’s exact test was used to determine whether there was a correlation between the gene status and the response to ICI treatment. If the *P* value was less than 0.05, H0 was rejected, meaning the gene status was considered related to the clinical response. **b,** The composition of the patient’s gene sentence. First, the WES genes of reference set were used to determine the candidate gene set based on a *P* value of less than 0.05 from the Fisher’s exact test. Then, the gene sentence for each patient in the reference set and test set was created by taking the intersection of all wild-type genes in the patient’s WES data and the candidate gene set. **c,** Predict responder and non-responder from the gene sentence. First, transform the gene sentences of the corpus into a heterogeneous graph with nodes representing either patients or genes, which establishes the relationship between different cohorts and patients with various tumor types. Information regarding to clinical response and gene mutation status is transmitted between patient nodes and gene nodes, so the probability of the response of a newly added patient can be predicted by integrating the existing heterogeneous information of the whole graph. Then, the cut off value can be determined to differentiate responders and non-responders by using the ROC curve of the reference set and the Youden’s index. After obtaining the patient grouping, the KM curve can then be plotted to evaluate the predictive performance. Abbreviations: WES, whole exome sequencing; R, responder; NR, non-responder; KM, Kaplan-Meier.

In constructing a gene sentence, gene words were chosen from the candidate gene set, as shown in Fig. 1b. If any of these genes underwent a mutation, they were excluded, leaving the remaining gene words to describe the patients’ genetic background. The maximum number of genes that can be included in a patient’s gene sentence is a hyperparameter *L*, which we have set to 468. If the gene sentence exceeds *L*, the candidate gene set is ordered ascendingly by the *P* value, and the top *L* genes are selected. We used the same candidate gene set and method to determine the gene sentence for the test set patients. To avoid the influence of artificially given gene order, we shuffle the gene words order after obtaining the gene sentence.

After collecting gene sentences, we use patients and genes as nodes to build graphs, as shown in Fig. 1c. The edges between genes are determined by PMI, while the edges between patient nodes and gene nodes are determined by IDF:

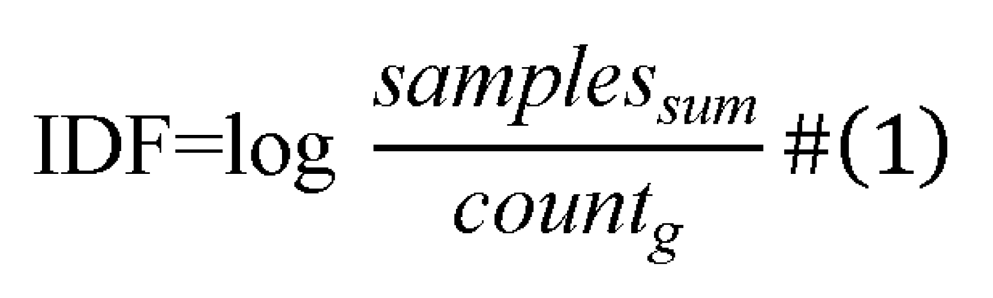

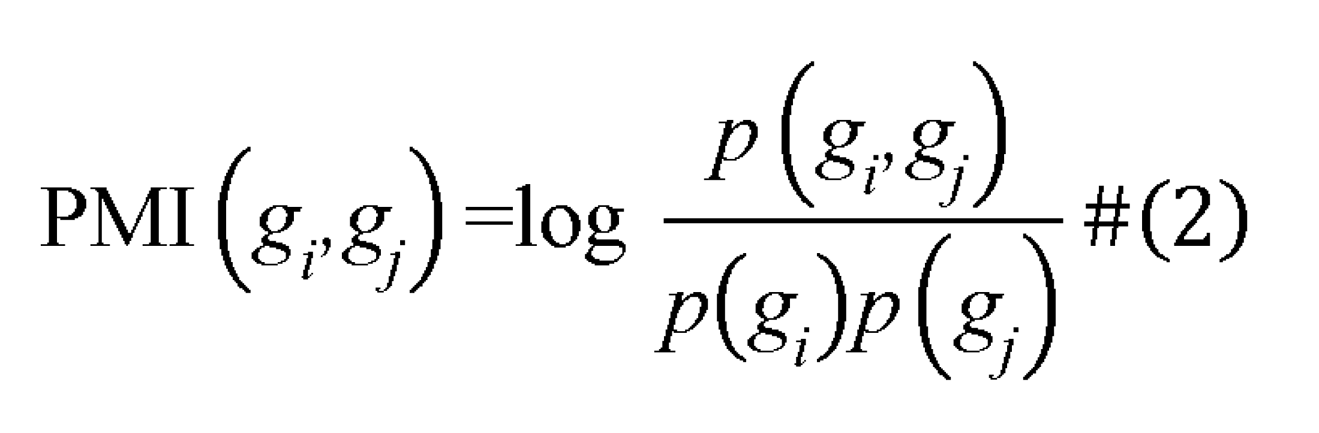

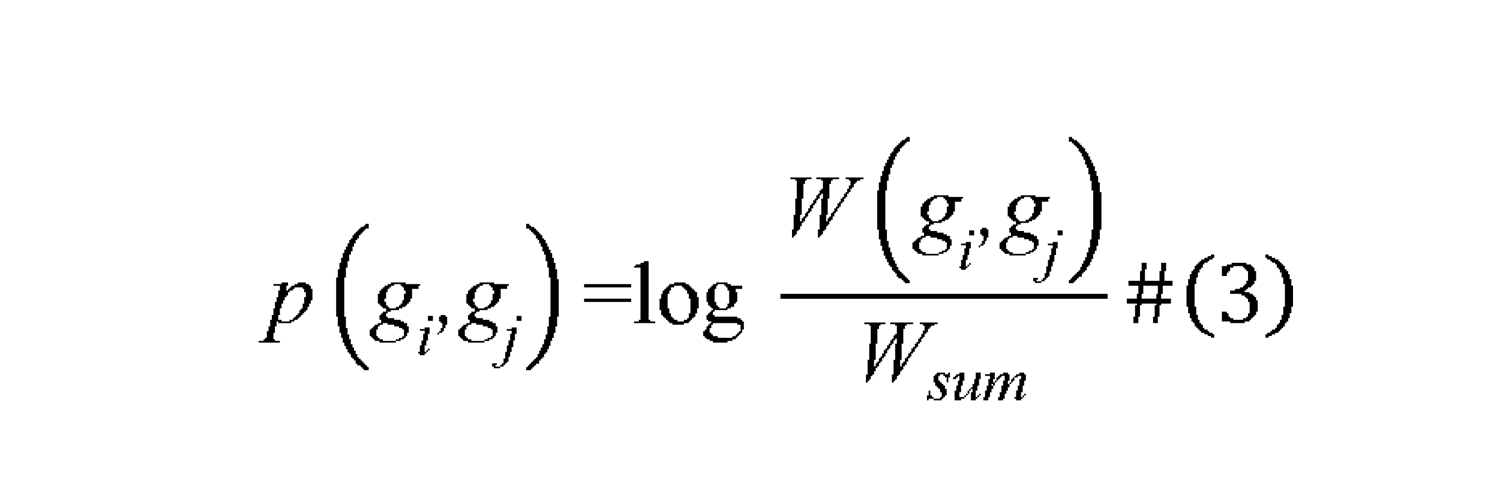

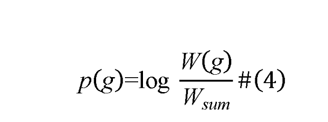

The variable *“Samples_sum_”* represents the total number of patients, *“count_g_”* represents the number of gene sentences containing a specific gene, and *“g”* represents any gene that appears in the gene sentences. The variable *“W_sum_”* represents the number of sliding windows for all gene sentences of the reference set and test set, *“W (g_i_,g_j_)”* represents the number of sliding windows where both genes *g_i_* and *g_j_* appear, and *“W(g)”* represents the number of sliding windows where a specific gene appears. We construct a graph that includes one test set at a time, along with the reference set. Therefore, the model is inherently transductive.

The sigmoid function was utilized to compress the output value of the final layer into the range of 0 to 1, which represents the probability of response to ICI treatment for each patient. Once the heterogeneous graph was constructed using the reference set, we randomly selected 80% of the reference set as the training set and the rest 20% as the validation set. Training was stopped when the loss on the validation set ceased to decrease, and we obtained the predicted response probability of ICI therapy for all patients in the reference and test sets. Subsequently, we plotted the ROC curve for the reference set and identified the decision threshold for responders and non-responders based on the Youden’s index that maximizes (Sensitivity + Specificity − 1)[17]. In the test set, patients with predicted probabilities greater than the threshold are considered responders, while those with predicted probabilities less than the threshold are considered non-responders. The predicted response classification for the test sets according to the decision threshold was then used for downstream analysis, such as KM curve plotting and comparison of TMB and neoantigen load (NAL).

### Materials

To analyze the feasibility of our method, we collected nine whole exome sequencing data for validation. The Miao2019 cohort comprises renal clear cell carcinoma patients treated with anti-PD-1 drugs [18]. The Hugo and Riaz cohorts consist of melanoma patients treated with anti-PD-1 drugs [19, 20]. The Miao2018 cohort comprises pan-cancer patients treated with either 1) anti-cytotoxic T lymphocyte-associated protein-4 (CTLA-4) drugs, 2) anti-PD-1 drugs, or 3) a combination of both anti-CTLA-4 and anti-PD-1 drugs [21]. The Rizvi cohort comprises non-small cell lung cancer (NSCLC) patients treated with anti-PD-1 drugs[22]. The Snyder and Van Allen cohorts consist of melanoma patients treated with anti-CTLA-4 drugs [23, 24]. These seven datasets were used as the reference set. The Hellmann cohort comprises non-small cell lung cancer patients treated with both anti-CTLA-4 and anti-PD-1 drugs [25]. The Liu cohort comprises melanoma patients treated with anti-PD-1 drugs [26]. These two datasets were designated as the test sets (see supplementary material Table S1). We downloaded eight WES datasets and corresponding clinical information from the cBioPortal database (https://www.cbioportal.org). The Riaz cohort was obtained from the original literature[21].

The collected data pertains to the drug response of cancer patients receiving ICI therapy, which has been filtered further. Firstly, we removed three tumor types with a sample size of less than 10. We also excluded 33 samples with non-evaluable response (NE), 7 samples that were not profiled, and 7 samples of “OTHER CONCURRENT THERAPY”. Additionally, we deleted 151 duplicate samples in the Miao2018 cohort. This cohort had 27 overlapping samples with Rizvi cohort, 37 with Snyder cohort, and 87 with Van Allen cohort.

Table 1 summarizes the characteristics of the filtered data. The reference set includes five tumor types: Melanoma (n=282), Non-Small Cell Lung Cancer (60), Renal Cell Carcinoma (35), Bladder Cancer (27), and Head and Neck Cancer (10). The test set contains two tumor types: Non-Small Cell Lung Cancer (68) and Melanoma (140). The reference set includes the following types of drug treatment: anti-PD-1 (166), anti-CTLA-4 (174), and anti-CTLA-4 + anti-PD-1 (74), whereas the test set includes anti-CTLA-4 + anti-PD-1 (68) and anti-PD-1 (140). The proportion of responders in the reference set is 27%. In the test set, it is 35% for the Hellmann cohort and 39% for the Liu cohort. The testing set of the Hellmann cohort does not contain any OS records.

**Table 1.** Characteristics of the reference set and two test sets.

| Characteristic | Num (Portion) |  |  |
| --- | --- | --- | --- |
|  | Reference set | Test set 1 (Hellmann cohort) | Test set 2 (Liu cohort) |
| **Gender** |  |  |  |
| Male | 220 (53%) | 32 (47%) | 82 (59%) |
| Female | 129 (31%) | 36 (53%) | 58 (41%) |
| NA | 65 (16%) | - | - |
| **Age** |  |  |  |
| <65 | 160 (39%) | 33 (49%) | - |
| >=65 | 105 (25%) | 35 (51%) | - |
| NA | 149 (36%) | - | 140 (100%) |
| **Cancer type** |  |  |  |
| Melanoma | 282 (68%) | - | 140 (100%) |
| Non-Small Cell Lung Cancer | 60 (15%) | 68 (100%) | - |
| Renal Cell Carcinoma | 35 (8%) | - | - |
| Bladder Cancer | 27 (7%) | - | - |
| Head and Neck Cancer | 10 (2%) | - | - |
| **Drug target** |  |  |  |
| anti-PD-1 | 166 (40%) | - | 140 (100%) |
| anti-CTLA-4 | 174 (42%) | - | - |
| anti-CTLA-4 + anti-PD-1 | 74 (18%) | 68 (100%) | - |
| **Response** |  |  |  |
| R (PR + CR) | 111 (27%) | 24 (35%) | 55 (39%) |
| NR (PD + SD) | 303 (73%) | 44 (65%) | 85 (61%) |
| **Overall patients** | 414 | 68 | 140 |

PR: partial response. CR: complete response. PD: progressive disease. SD: stable disease. NA: not available.

In addition to drug response data, we collected the data from TCGA to further validate our model, including the WES data, RNA-seq data, and patients’ overall survival time. This allowed us to examine the distribution of known key gene signatures across different predicted response groups. The WES and RNA-seq data were obtained using TCGAbiolinks [27], and the survival time data were collected from Liu et al [28]. Additionally, we obtained Cibersort immune infiltration values for each TCGA cancer sample from Thorsson et al [29].

### GSEA analysis

We used the whole ICI therapy datasets to train TG468 and predicted TCGA datasets. Following the process shown in Fig. 1, we predicted the probabilities of the TCGA datasets using TG468 and identified the decision threshold for the whole ICI therapy datasets using Youden’s index. Additionally, we divided the TCGA datasets into R and NR groups based on the decision threshold. To analyze the prediction results, we used TCGA RNA-seq data to identify differentially expressed genes using the DESeq2 package [30]. We then performed GSEA analysis based on the Kyoto Encyclopedia of Genes and Genomes (KEGG) pathway using the clusterProfiler package [31]. Gene sets with an FDR (Benjamini-Hochberg method) lower than 0.25 were considered significantly enriched.

### Statistical analysis

We used the two-sided Fisher exact test to select candidate gene sets. It should be noted that we did not perform multiple hypothesis testing correction here, which would perform better in controlling false positives but would also filter out many potential candidate genes. Accordingly, we transformed the problem of further feature selection and combination optimization into a learning task based on Text GCN. We used the two-sided Wilcoxon rank sum test to compare the TMB and NAL of ICI therapy data as well as TCGA gene expression levels between predicted responders and non-responders. Additionally, we plotted KM curve of PFS and OS using the logrank test based on predicted groups, which used χ2 test statistic to calculate *P* values[32]. Fisher’s exact test was implemented by python package scipy[33]. Logrank test and Cox Proportional-Hazards analysis and Wilcoxon rank sum test were implemented by the survminer package and ggsignif package in R version 4.1.2 (https://www.r-project.org).

## RESULTS

### Evaluation of TG468 as a biomarker for response to ICIs

The AUC-ROC of the reference and test set 1 (Hellmann cohort) are 0.77 and 0.71, respectively (Fig. 2a). The AUC-ROC of the reference and test set 2 (Liu cohort) are 0.87 and 0.61, respectively (Fig. 2c). The difference in AUC for the reference set may be due to the different test sets used in constructing the heterogeneous graphs. Using Youden’s index, cutoff values for responders and non-responders were determined based on the ROC curve of the reference set, resulting in cutoff values of 0.257 and 0.206 for the two graphs, respectively. Any predicted values greater than the cutoff values are classified as responders, while values less than the cutoff values are classified as non-responders.

**Fig. 2.**
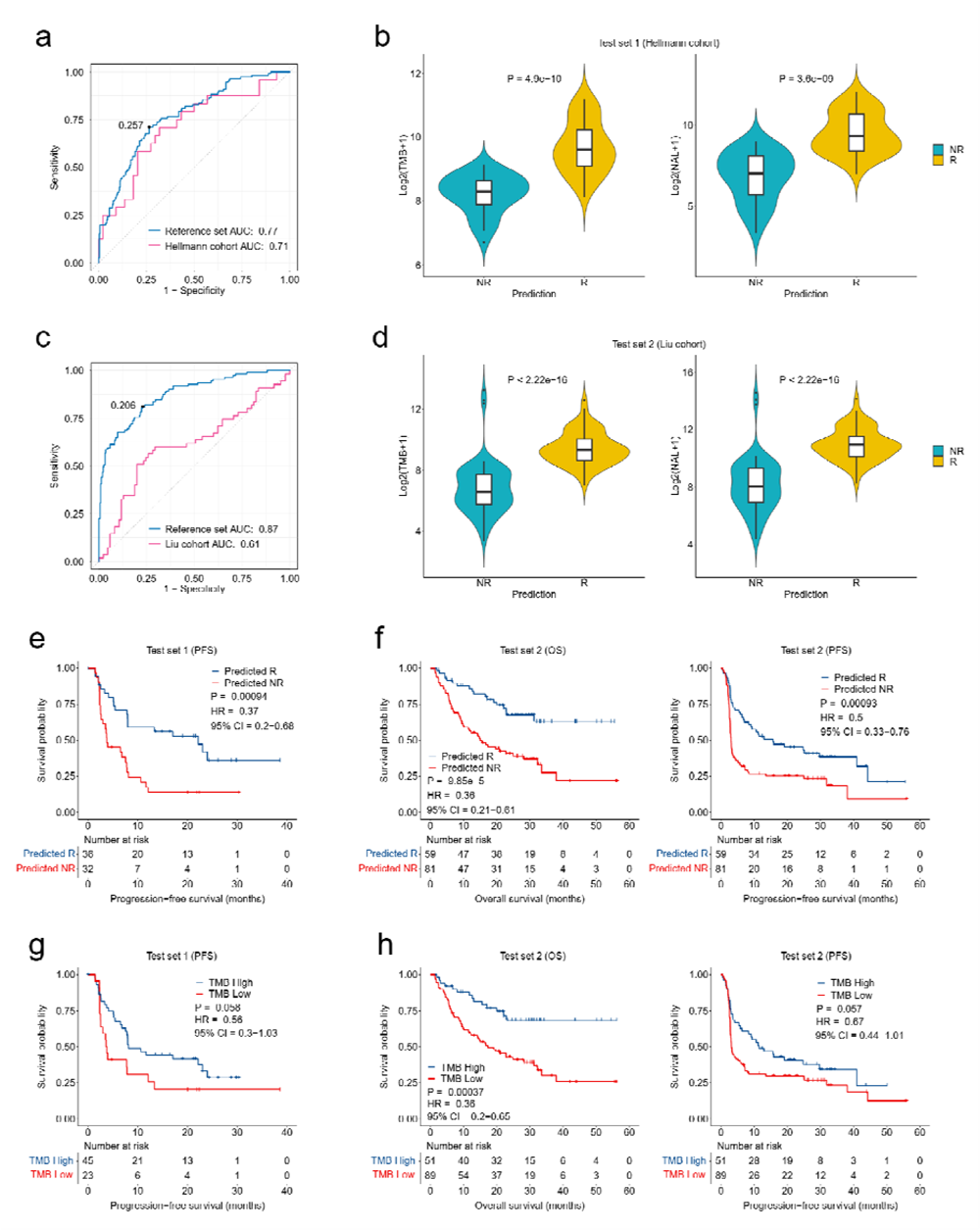
Assessment of TG468’s predictive performance. **a,** ROC curves of TG468 on the reference set and test set 1 (Hellmann cohort). According to the reference set’s ROC curve and Youden’s index, the cutoff is determined to be 0.257 for dividing responders and non-responders. **b,** Differences of TMB and NAL between R and NR groupings on the test set 1. **c,** ROC curves of TG468 on the reference set and test set 2 (Liu cohort). According to the reference set’s ROC curve and Youden’s index, the cutoff is determined to be 0.206 for dividing responders and non-responders. **d,** Differences of TMB and NAL between R and NR groupings on the test 2. **e,** KM curve of PFS based on the TG468’s prediction results on test set 1. **f,** KM curves of OS and PFS based on the TG468’s prediction results on test set 2. **g,** KM curve of PFS based on the TMB (10 mut/Mb) cutoff groupings on test set 1. **h,** KM curves of OS and PFS based on the TMB (10 mut/Mb) cutoff groupings on test 2. Abbreviations: TMB, tumor mutational burden; NAL, neoantigen load; R, responder; NR, non-responder; KM, Kaplan-Meier; OS, overall survival; PFS, progression-free survival; HR, hazard ratio; CI, confidence interval.

Fig. 2b and 2d show significant differences between responders and non-responders in terms of TMB and NAL in the two test sets. The higher TMB and NAL in the R group indicate higher immunogenicity, suggesting a greater likelihood of clinical benefit after receiving ICI therapy. To demonstrate the prediction performance of TG468, KM curves for OS and PFS were plotted using the predicted results. Fig. 2e displays the PFS KM curves for test set 1 (Hellmann cohort). The PFS for the predicted R group was significantly longer than that for the predicted NR group (P = 0.00094, hazard ratio [HR] = 0.37, 95%CI = 0.2-0.68). Fig. 2f shows the OS and PFS KM curves for test set 2 (Liu cohort), respectively. OS for the predicted R group was significantly longer than that for the predicted NR group (P = 9.85e-5, HR = 0.36, 95%CI = 0.21-0.61). Additionally, PFS for the predicted R group was significantly longer than that for the predicted NR group (P = 0.00093, HR = 0.5, 95%CI = 0.33-0.76). These results demonstrate that TG468 is a powerful tool for selecting patients who are likely to benefit from ICI therapy.

### Comparison of TG468 with TMB for response to ICIs

For comparison, we also evaluated tumor mutational burden (TMB) as a potential biomarker for predicting outcomes of immune checkpoint inhibitor (ICI) therapy in the two test sets. In this analysis, we used non-synonymous mutation counts as the measure of TMB, with a cutoff value of 10 mut/Mb [5]. Fig. 2g (P = 0.058, HR = 0.56, 95%CI = 0.3-1.03) shows the progression-free survival (PFS) Kaplan-Meier (KM) curves for the test set 1 (Hellmann cohort) based on different TMB groupings. Fig. 2h shows the overall survival (OS) and PFS KM curves for the test set 2 (Liu cohort) based on different TMB groupings (OS: P = 0.00037, HR = 0.36, 95%CI = 0.2-0.65; PFS: P = 0.057, HR = 0.67, 95%CI = 0.44-1.01). As a predictive biomarker, the *P* values of KM curves plotted based on TMB groupings for both test sets are greater than 0.05. Although TMB can distinguish PFS of patients to a certain extent, its predictive performance for PFS is limited. We can see that compared with TMB, the *P* values of the OS and PFS KM curves plotted by TG468 prediction results are significantly smaller, and the HR coefficient for PFS is lower.

Note that TMB cutoffs have been defined differently across studies and in various patient populations. In our study, we also used the top 25% of TMB as the cutoff value for high TMB and low TMB [34, 35]. Supplementary material Fig. S1-a shows PFS KM curves of test set 1 according to TMB (top 25%) (P = 0.0027, HR = 0.31, 95%CI = 0.14-0.69), and supplementary material Fig. S1-b shows the OS and PFS KM curves of the test set 2 according to TMB (top 25%) (OS: P = 0.0043, HR = 0.37, 95%CI = 0.19-0.75; PFS: P = 0.32, HR = 0.79, 95%CI = 0.49-1.26). We may find that, compared with TMB (top 25%), the *P* values of the OS and PFS KM curves plotted by TG468 prediction results is still significantly smaller.

Supplementary material Fig. S2 compares TG468 and TMB in terms of positive predictive value (PPV), negative predictive value (NPV), sensitivity, specificity, and accuracy. In the Hellmann cohort, TG468 outperformed TMB (top 25%) on two performance metrics: PPV and specificity. Additionally, TG468 outperformed the TMB (10mut/Mb) cutoff on three metrics, excluding PPV and specificity. In the Liu cohort, TG468 outperformed both the TMB (top 25%) and TMB (10mut/Mb) cutoffs on four metrics, excluding NPV. Overall, these results demonstrate that TG468’s predictive performance is superior to that of TMB across various tumor types.

### Comparison of TG468 with single-gene biomarkers for response to ICIs

To compare with single-gene biomarkers, we collected a list of single gene biomarkers that have been reported to be predictive of the response to ICI therapy. These biomarkers include ARID1A [36], EPHA7[3], KEAP1[37], KMT2 family[38], LRP1B[39], MGA[40], MUC16[41], NOTCH4[42], PAPPA2[2], PBRM1[12], POLD1[43], POLE[44], and TTN[13]. We used a univariate Cox proportional hazards regression model to analyze the correlation between mutation gene or predicted responders and PFS. The results of this analysis are shown in Fig. 3a and 3b. In the Hellmann cohort, LRP1B (HR, 0.45 [95%CI, 0.22-0.92], P = 0.028), MGA (HR, 0.31 [95%CI, 0.11-0.87], P = 0.026), PAPPA2 (HR, 0.40 [95%CI, 0.17-0.96], P = 0.039), and TTN (HR, 0.50 [95%CI, 0.27-0.92], P = 0.026) single-gene biomarkers were able to predict the clinical benefit of ICI therapy. Compared with these single-gene biomarkers, TG468’s prediction (HR, 0.37 [95%CI, 0.20-0.68], P = 0.001) showed the second lowest risk of progression, only surpassed by MGA. On the Liu cohort, only TG468’s prediction showed the ability to predict clinical benefit of ICIs (HR, 0.51 [95%CI, 0.33-0.77], P = 0.001), while all other single-gene biomarkers did not.

**Fig. 3.**
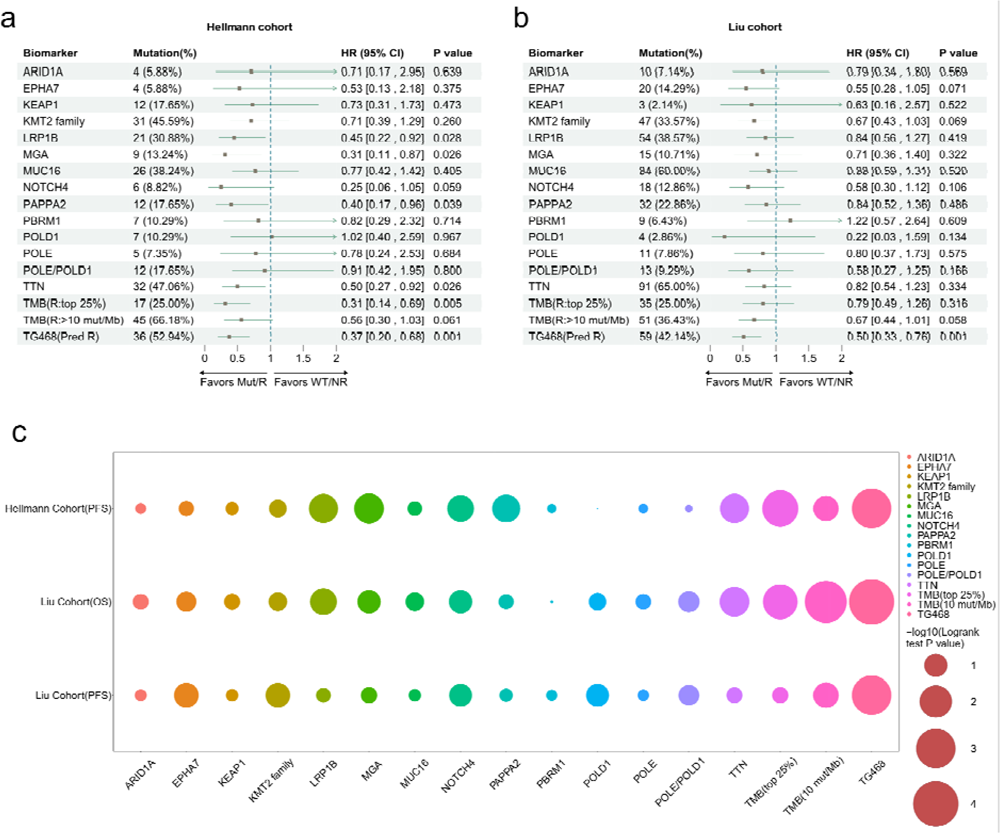
Comparison of the predictive performance of single-gene biomarkers and TMB with TG468. **a,** Comparison using univariable Cox proportional hazards regression analysis on the Hellmann cohort. **b,** Comparison using univariable Cox proportional hazards regression analysis on the Liu cohort. **c,** Comparison of biomarkers with corresponding *P* values of KM curves after taking the negative logarithm operation using the logrank test. Abbreviations: HR, hazard ratio; CI, confidence interval; KM, Kaplan-Meier.

In addition, KM curves were plotted, and logrank test *P* values were used to evaluate the predictive performance of single-gene biomarkers based on their mutation status. Fig. 3c displays these values after taking the negative logarithm. Each column bubble of the same color represents a biomarker’s *P* values on the two test sets. The larger the circle, the smaller the *P* value, and the better the prediction. In comparison, TG468 exhibited superior prediction results to those of single-gene biomarkers.

As shown in Fig. 3a and 3b, although TMB (top 25%) showed a lower risk of progression (HR, 0.31 [95% CI, 0.14-0.69], P = 0.005) than TG468’s prediction (HR, 0.37 [95% CI, 0.20-0.68], P = 0.001) on the Hellmann cohort, TG468’s prediction was still able to predict clinical benefit of ICI therapy on the Liu cohort (HR, 0.50 [95% CI, 0.33-0.76], P = 0.001) whereas TMB (top 25%) failed (HR, 0.79 [95% CI, 0.49-1.26], P = 0.316). Moreover, on both tests, TG468’s prediction exhibited greater ability to predict clinical benefit of ICIs than TMB (10 mut/Mb) (Hellmann cohort: HR, 0.56 [95% CI, 0.30-1.03], P = 0.061; Liu cohort: HR, 0.67 [95% CI, 0.44-1.01], P = 0.058). In addition, TG468’s prediction yielded more significant logrank test *P* values in survival analysis than TMB (top 25%) and TMB (10 mut/Mb), as shown in Fig. 3c.

TMB with different thresholds and specific single-gene biomarkers may show lower risk of progression in specific cohorts, such as MGA and TMB (top 25%) in the above analyses, but may not perform well on all test sets. These results suggest that these biomarkers may only effectively predict progression risk in specific tumor types, rather than across all types of tumors. However, TG468 showed the ability to predict progression risk in all test sets. When comparing logrank *P* values obtained from survival analyses of different biomarkers, TG468 showed more significant *P* values than other biomarkers in predicting progression-free and overall survival. In summary, TG468 demonstrated greater generalization ability to predict the risk of progression across tumor types and has better survival prediction ability compared to single-gene biomarkers and TMB. This suggests that TG468 may be a promising biomarker for predicting response to ICIs.

### Potential mechanisms of TG468’s association with antitumor immunity

To explain why TG468 is effective in predicting the clinical benefit of ICI treatment, we investigated its potential mechanisms associated with antitumor immunity. We applied TG468 to predict TCGA data and analyzed the relationship between the predicted grouping and key biological signatures, including its association with the expression of immune-related genes and the level of immune cell infiltration. These analyses can verify whether TG468’s prediction is related to the biological signatures that have been reported to affect ICI treatment, and guide the precise use of ICI in clinical practice.

#### Assessment of TG468’s immune status stratification and prognostic role in SKCM patients

TG468 was trained using 9 ICI therapy datasets, and then the model was used to analyze TCGA WES data for patient stratification and prognostic analysis. As the largest proportion of the 9 datasets came from melanoma patients, we first used TG468 to stratify TCGA-skin cutaneous melanoma (SKCM) patients. Downstream analysis included examining survival differences and immune status differences among different predicted groups, as well as conducting GSEA analysis to further validate the rationality of the model’s predictions.

The results in Fig. 4a show a time-dependent ROC curve based on the prediction. It is observed that the prediction performance for survival time of 1-year, 3-year, and 5-year is similar, all around 0.6. After determining the threshold for dividing patients into R and NR groups based on the 9 ICI therapy data sets, TCGA-SKCM patients were divided into R and NR groups accordingly. In Fig.4b, the KM curve based on the predicted groupings shows that the patients in the R group have a significantly longer survival time than those in the NR group (P = 0.0018, HR = 0.65, 95%CI = 0.50-0.86). These results demonstrate that our model can predict the prognosis of TCGA-SKCM patients.

**Fig. 4.**
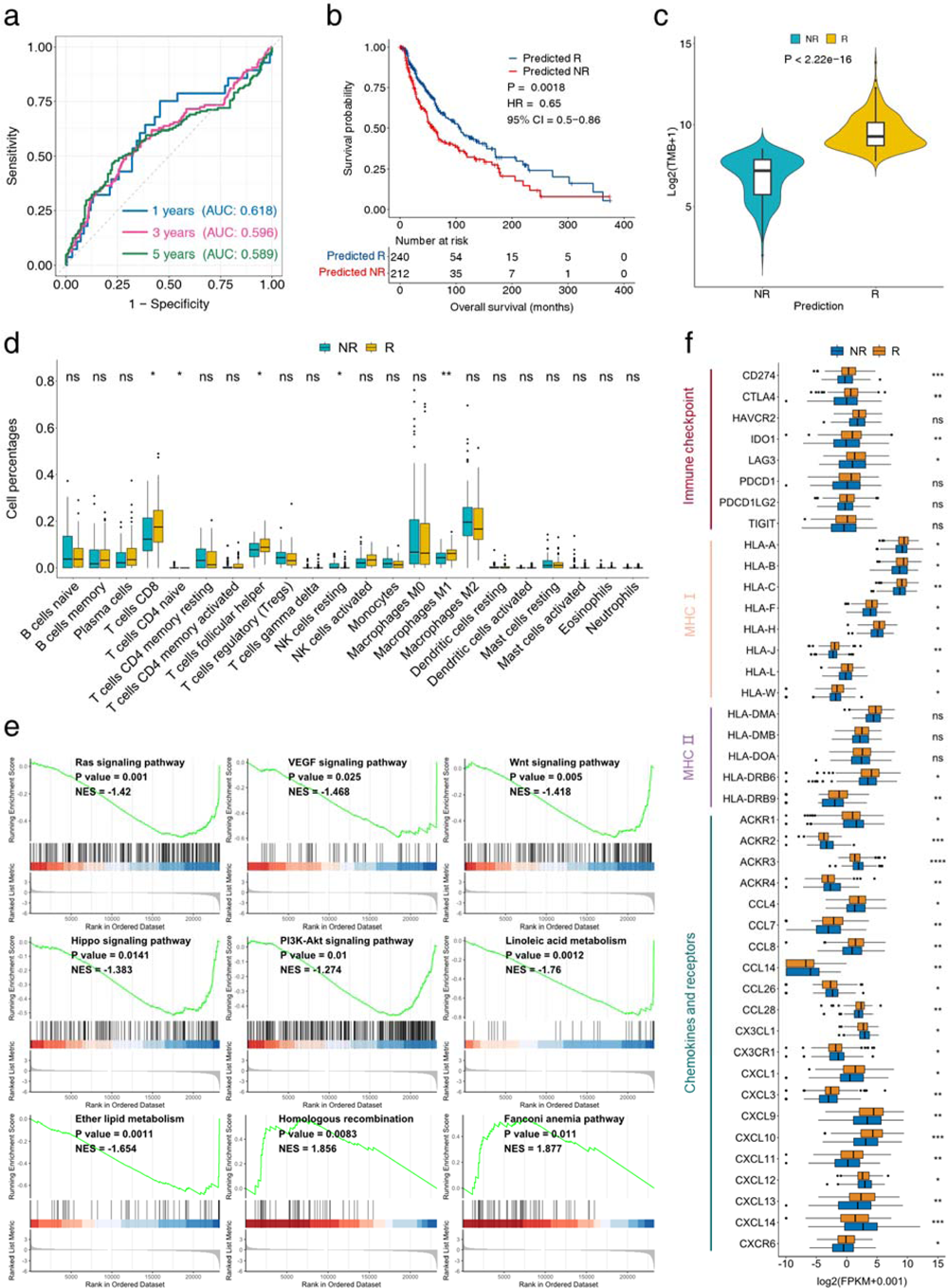
Analysis of the prediction results of TG468 in SKCM. **a.** Time-dependent ROC curve based on the predicted response probability of SKCM. **b.** OS KM curve based on the predicted grouping of SKCM. **c.** Infiltration analysis of immune cells based on the predicted grouping of SKCM. **d.** Results of gene set enrichment analysis (FDR< 0.25) based on the predicted grouping of SKCM. **e.** Analysis of immune-related gene expression based on the predicted grouping of SKCM. \**P* < 0.05, \*\**P* < 0.01, \*\*\**P* < 0.001, \*\*\*\**P* < 0.0001. Abbreviations: SKCM: skin cutaneous melanoma, OS: overall survival, KM: Kaplan-Meier, GSEA: gene set enrichment analysis.

Fig. 4c shows that the predicted R group with higher TMB values suggests a higher likelihood of response to ICI therapy due to increased immunogenicity. Cibersort analysis was conducted based on predicted groupings, revealing that the composition of immune cells differed significantly between the R and NR groups (Fig. 4d). Immune infiltration analysis showed that the tumor microenvironment of the R group had significantly higher levels of CD8 T cells and macrophages m1 cells compared to the NR group. This is consistent with previous reports stating that CD8 T cells play a direct role in tumor cell killing and are key determinants of response to ICIs[45]. Macrophage m1 cells are also key tumor suppressor cells that are related to T cell stimulation and ICI therapy [46, 47].

Furthermore, based on the predicted grouping results, GSEA analysis was performed, as shown in Fig. 4e. The Ras signaling pathway, vascular endothelial growth factor (VEGF) signaling pathway, Wnt signaling pathway, hippo signaling pathway, PI3K-Akt signaling pathway, linoleic acid metabolism pathway, and ether lipid metabolism pathway were upregulated in the predicted NR group. Meanwhile, the homologous recombination pathway and fanconi anemia pathway were upregulated in the predicted R group. These pathways have previously been reported to affect ICI therapy, which are consistent with current findings. Activation of the Ras signaling pathway leads to an immunosuppressive tumor microenvironment, hindering the activation and infiltration of T cells and affecting the therapeutic effect of ICIs[48]. VEGF can directly affect and promote the survival, proliferation, and invasion of tumor cells, while inhibiting the function of T cells, contributing to the immunosuppressive microenvironment[49, 50], and potentially affecting the efficacy of ICI treatment. The Wnt signaling pathway is associated with ICIs treatment response, driving immune cell exclusion from the tumor microenvironment, and may be a driving factor of immunotherapy drug resistance[51]. The hippo signaling pathway and PI3K-Akt signaling pathway are associated with ICIs resistance[52]. Increased lipid content correlates with ICI response[53], and linoleic and ether lipid accumulation may imply a greater likelihood of response to ICI therapy. The absence of DNA damage repair (DDR) pathways may promote tumor progression, and DDR pathways are associated with the response of ICIs treatment [54, 55].

Furthermore, as illustrated in Fig. 4f, various immune checkpoint-related genes (CD274, CTLA4, IDO1, LAG3) showed significantly higher expression in the R group. The MHC1 and MHC2 genes also exhibited significantly higher expression in the R group, while chemokine and receptor molecules demonstrated significantly different expression levels between the R and NR groups. Previous studies have suggested that higher expression of immune checkpoint-related genes is indicative of a better response to ICIs therapy[56]. In tumor tissues, MHC1 is responsible for presenting antigens to CD8 T cells, and its down-regulation is associated with resistance to ICIs[57, 58]. Additionally, MHC2 positive expression correlates with ICIs therapy response[59]. CXCL9, CXCL10, CXCL11 and CXLC13 also showed significantly higher expression in the R group, which is consistent with previous findings. Increased expression of CXCL9, CXCL10 and CXCL11 can enhance T cell infiltration and promote the therapeutic effect of ICIs[60-62]. CXCL13 expression can generate effector T cells and is also associated with ICIs treatment response[63].

These findings indicate that TG468 is effective in distinguishing the immune status of SKCM patients and can justify the model’s predictions on ICI therapy response. The R group exhibited significantly higher expression levels of immune cell infiltration, immune checkpoint-related genes, antigen presentation-related genes, and certain T-cell infiltration-related chemokines, which were associated with clinical benefits after receiving ICIs treatment. GSEA analysis demonstrated that enriched pathways are related to the response to ICIs therapy. These findings suggest that the prediction of TG468 for ICIs therapy response is reasonable. Furthermore, the difference in immune cell infiltration between the R and NR groups may also explain the ability of TG468 to predict prognosis. For instance, a higher level of CD8 T cells in the tumor microenvironment leads to better clinical outcomes for patients [64], because they are responsible for killing tumor cells. This may be the reason why the R group has a significantly longer survival time than the NR group.

#### Assessment of TG468’s immune status stratification and prognostic role in pan-cancer patients

TG468 utilizes data from various tumor types to construct a heterogeneous graph, which suggests that the model may have learned common molecular characteristics that can indicate the response of different tumor types to ICI therapy. Therefore, we used TG468 to stratify pan-cancer patients in TCGA, and we are interested in whether there are significant differences in the distribution of known key biological signatures related to ICI among different prediction groups. This result may indirectly indicate whether TG468 has learned molecular commonalities behind the response of different tumor types to ICI treatment. The collection of 9 ICI treatment datasets was used to train TG468, then the model was used for patient stratification and prognostic analysis of different cancer types. Based on the predicted grouping, differences in the level of immune cell infiltration and the expression of genes related to immune checkpoints were analyzed. Subsequently, GSEA was conducted on TCGA datasets, revealing alterations in immune-related pathways.

As shown in Fig. 5 a-b and Fig. S4 a-e, there are significant differences in the immune microenvironment cell composition of several cancer types, including Uterine Corpus Endometrial Carcinoma (UCEC), Cervical squamous cell carcinoma and endocervical adenocarcinoma (CESC), Lung adenocarcinoma (LUAD), Bladder Urothelial Carcinoma (BLCA), Colon adenocarcinoma (COAD), Lung squamous cell carcinoma (LUSC) Breast invasive carcinoma (BRCA). The composition of CD8 T cells and/or activated CD4 memory T cells in predicted responder (R) group was significantly higher than that of predicted non-responder (NR) group. Except for LUSC, immune checkpoint-related genes were significantly more highly expressed in the R group compared to the NR group. Studies have shown that patients with better immune infiltration (CD8/CD4 T cells) and higher expression of immune checkpoint-related genes are more likely to benefit clinically from ICIs treatment[45, 56, 65-67].

**Fig. 5.**
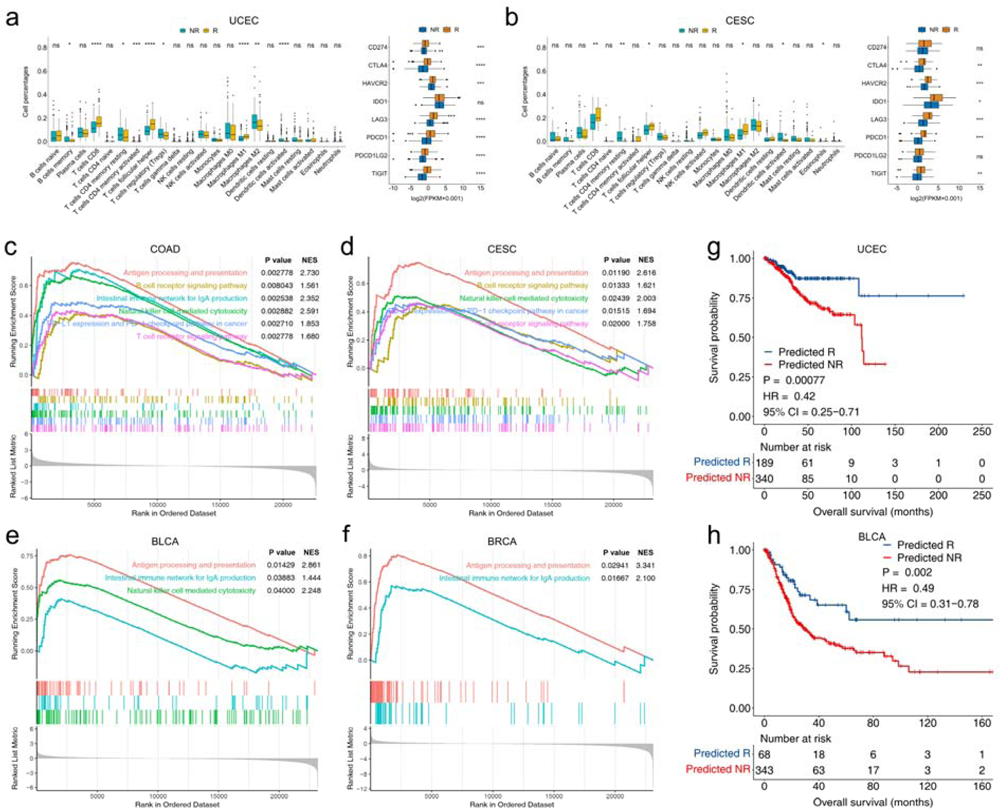
Analysis of TG468’s prediction results on pan-cancer datasets. **a-b,** Immune infiltration analysis and immune checkpoint-related gene expression analysis based on prediction results of UCEC and CESC (\**P* < 0.05, \*\**P* < 0.01, \*\*\**P* < 0.001, \*\*\*\**P* < 0.0001). **c,** GSEA analysis of COAD reveals significant enrichment in six immune-related pathways. **j,** GSEA analysis of CESC reveals significant enrichment in five immune-related pathways. **k,** GSEA analysis of BLCA reveals significant enrichment in three immune-related pathways. **l,** GSEA analysis of BRCA reveals significant enrichment in two immune-related pathways. **m,** UCEC patients classified as responders have a significantly longer overall survival compared to non-responders. **n,** BLCA patients classified as responders have a significantly longer overall survival compared to non-responders. Abbreviations: GSEA, gene set enrichment analysis (FDR< 0.25); UCEC, Uterine Corpus Endometrial Carcinoma; CESC, Cervical squamous cell carcinoma and endocervical adenocarcinoma; COAD, Colon adenocarcinoma; BLCA, Bladder Urothelial Carcinoma; BRCA, Breast invasive carcinoma.

Further GSEA analysis was performed on the seven types of TCGA data mentioned above, revealing that immune-related pathways were enriched in COAD, CESC, BLCA and BRCA, as shown in Fig. 5 c-f. Among them, six different immune pathways related to the response of immune therapy were enriched in the predicted R group. The antigen processing and presentation pathway was enriched in the predicted R group, consistent with the report that gene expression changes related to antigen processing and presentation are associated with antigen presentation and may influence the response to ICIs therapy [68]. The B Cell Receptor (BCR) signaling pathway was upregulated in the predicted R group, which is associated with ICIs treatment response[69] as BCR signaling affects B cells survival and B cells can predict response to ICI treatment[70, 71]. The intestinal immune network for IgA production pathway was upregulated in the predicted R group, in line with the association between intestinal IgA production and host-microbiota interactions, as gut microbiota can regulate the body’s immune function and optimize the therapeutic effect of immune checkpoint inhibitors[72-74]. The natural killer cell mediated cytotoxicity pathway was upregulated in the predicted R group, indicating that the use of ICIs will make the tumor-killing process more efficient by restoring the antitumor cytotoxic activity of NK cells[75]. PD-L1 expression and PD-1 checkpoint pathway in cancer were upregulated in the predicted R group, consistent with the association between PD-L1 expression and response to immunotherapy, and the fact that PD-1, as one of the targets of ICI inhibitors, is directly related to ICI response [76, 77]. The T cell receptor signaling pathway was also upregulated in the predicted R group, consistent with its role in T cell activation and development [78].

Furthermore, TG468 predicted that responders had a significantly longer overall survival time than non-responders in UCEC and BLCA, as depicted in Fig. 5g and 5h. This observation may be attributed to the higher levels of immune infiltration in the responder group. The model’s predictive ability for ICI therapy response was further validated by the observation of elevated levels of T cell infiltration, increased expression of immune checkpoint genes, and upregulation of immune pathways, which were associated with a higher probability of response to ICI therapy. This validation was observed across different tumor types, including UCEC, CESC, COAD, and BRCA, which were not present in the ICI therapy dataset. Therefore, these results indicate that the model may have learned molecular signatures that can indicate the response to ICI therapy across various types of tumors.

## DISCUSSION

We developed the TG468 model to predict the probability of a patient’s response to immunotherapy. In the test sets, we found significant differences in TMB and NAL between the R and NR groups, with higher TMB and NAL values in the R group indicating higher immunogenicity. We evaluated the model’s performance using KM survival curves across different test sets. The KM curves for OS and PFS were more significant for TG468 predictions compared to TMB and single-gene biomarkers. Additionally, we compared the prediction performance of TG468 and TMB on test sets in terms of a variety of metrics. TG468 had better prediction performance than TMB. These findings demonstrate the superior predictive ability of TG468 compared to single-gene biomarkers and TMB, indicating its potential as a robust and effective tool for predicting immunotherapy response in different cancer patient populations. To achieve clinical response prediction for immunotherapy, sequencing of up to 983 genes is required for TG468. However, this offers the possibility of economic and rapid clinical application in the future.

There are several potential reasons why the model achieved better prediction performance than single-gene biomarkers and TMB. First, we selected a candidate gene set for the WES genes since the number of genes sequenced by WES is far greater than the number of patients in the entire dataset. This process avoids over-fitting of the model caused by the too-long gene sentences of some patients. Second, most somatic alterations identified by WES are likely passenger events that have no impact on tumors[79]. The status of passenger genes may not significantly affect ICIs therapy response, and the potential interference of these genes can be mitigated by the selection of candidate gene sets. Third, TG468 effectively integrates the impact of multiple biomarkers on patients as a whole biomarker to indicate response to ICI therapy. Moreover, the model connects patients in the same or different cohorts through gene nodes, allowing for discovering molecular commonalities among cancer patients across diseases. This could be the reason for accurately predicting response to ICI therapy.

To explain the potential mechanisms of TG468’s association with antitumor immunity, we analyzed TCGA data and investigated the relationship between key biological signatures that affect the response to ICI treatment and the prediction results. We first analyzed TG468’s prediction results for TCGA-SKCM data, as melanoma patients represent the largest proportion of ICI therapy data. We conducted immune infiltration analysis, GSEA analysis, and differential expression analysis of immune-related genes for the classification results. The immune infiltration analysis showed significant differences in the tumor microenvironment between the R and NR groups. The GSEA results were enriched in ICI therapy response-related pathways, and there were also significant differences in immune-related genes. Overall, these analyses demonstrate that the model can distinguish patients with different immune statuses and gene expressions in SKCM, and provide a potential explanation for the model’s prediction results. For example, the higher level of CD8 T cells in the tumor microenvironment of the R group compared to the NR group may indicate that patients in the R group are more likely to achieve a clinical response after using immune inhibitors. CD8 T cells play a pivotal role in determining the response to ICIs[45], in other words, PD-1 and CTLA-4 blockades are more effective in tumors infiltrated by CD8 T cells[80, 81].

To investigate whether TG468 has learned the molecular commonality behind the response of different tumor types to ICI therapy, we further analyzed its prediction results in TCGA data across 32 cancer types. We identified significant differences in immune infiltration and immune checkpoint-related gene expression among seven cancer types and six cancer types, respectively. Additionally, we conducted GSEA analysis of these seven tumors and found upregulation of immune-related pathways in the R group. These findings highlight the model’s ability to capture molecular commonalities across diseases that are indicative of differences in immune cell infiltration, immune checkpoint-related gene expression profiles, and immune pathway activation. Consequently, TG468 may serve as a valuable tool for identifying patients who could potentially benefit from ICI therapy, even in tumor types that were not included in the model’s training set. For example, in UCEC, CD8 T cells in the R group were significantly higher than those in the NR group, and PDCD1 and CTLA4 were significantly more highly expressed in the R group. Therefore, it is reasonable to recommend ICI therapy for patients in the R group.

A limitation of our study is that the model’s performance was only tested on two immunotherapy test sets. In the future, with access to more clinical experimental data, our method can be used to construct models with greater generalization ability, making it more suitable for rapid clinical application.

## CONCLUSION

TextGCN is a graph convolutional network that was originally proposed for text classification problems due to its excellent text representation capabilities. In this study, we use TextGCN to model the relationship between clinical response and the global co-occurrence sequence of gene status, formulating ICI response prediction as a text classification problem. The resulting model, TG468, predicts ICI therapy response and outperforms single-gene biomarkers and TMB in terms of predictive performance. Moreover, TG468 may have predictive power for ICI therapy response for tumor types not present in the training set, demonstrating its generalization capability. Our approach of using WES data as input for a model designed for NLP tasks represents a novel paradigm that may inspire future research. This paradigm opens up new possibilities for leveraging NLP techniques to analyze WES genomic data, paving the way for further advancements in the field of precision medicine.

## SUPPLEMENTARY MATERIAL

**Table S1** Description of WES Data sets that received ICI therapy. **Fig. S1** KM curves of the test sets based on the TMB (25%) cutoff groupings. **Fig. S2** Comparisons between TG468 and TMB were made on PPV, NPV, sensitivity, specificity, accuracy. **Fig. S3** Analysis of TG468’s prediction results on pan-cancer datasets.

## AUTHORS’ CONTRIBUTIONS

Kun wang conceived of and designed the study and drafted the manuscript. Jiangshan Shi accessed and verified the underlying data. Xiaochu Tong, Ning Qu, Xiangtai Kong, Shengkun Ni and Jing Xing participated in the analysis of the experimental results. Xutong Li and Mingyue Zheng provided administrative support and supervised the project. Mingyue Zheng Revised the manuscript. All authors read and approved the final version of the manuscript and are accountable for all aspects of the work. The authors declare that they have no competing interests.

## Data Availability

All data produced in the present study are available upon reasonable request to the authors

