## SUPPLEMENTARY MATERIAL for "TG468: A Text Graph Convolutional Network for Predicting Clinical Response to Immune Checkpoint Inhibitor Therapy"

Additional file

### Table of Contents

**Table S1** Description of WES Data sets that received ICI therapy. S3

**Fig. S1** KM curves of the test sets based on the TMB (25%) cutoff groupings. S3

**Fig. S2** Comparisons between TG468 and TMB were made on PPV, NPV, sensitivity, specificity, accuracy. S4

**Fig. S3** Analysis of TG468's prediction results on pan-cancer datasets. S5

**Table S1 Description of WES Data sets that received ICI therapy.**

| Data sets | Target | Cancer type | Patient number |
|---|---|---|---|
| Miao2019 cohort | Anti-PD-1 | Renal Clear Cell Carcinoma | 35 |
| Hugo cohort | Anti-PD-1 | Melanoma | 38 |
| Miao2018 cohort | Anti-CTLA-4<br>Anti-PD-1<br>Anti-CTLA-4 + Anti-PD-1 | Non-Small Cell Lung Cancer<br>Bladder Cancer<br>Melanoma<br>Head and Neck Cancer | 249 |
| Rizvi cohort | Anti-PD-1 | Non-Small Cell Lung Cancer | 35 |
| Snyder cohort | Anti-CTLA-4 | Melanoma | 64 |
| Van Allen cohort | Anti-CTLA-4 | Melanoma | 110 |
| Riaz cohort | Anti-PD-1 | Melanoma | 73 |
| Hellmann cohort | Anti-CTLA-4 + Anti-PD-1 | Non-Small Cell Lung Cancer | 75 |
| Liu cohort | Anti-PD-1 | Melanoma | 144 |

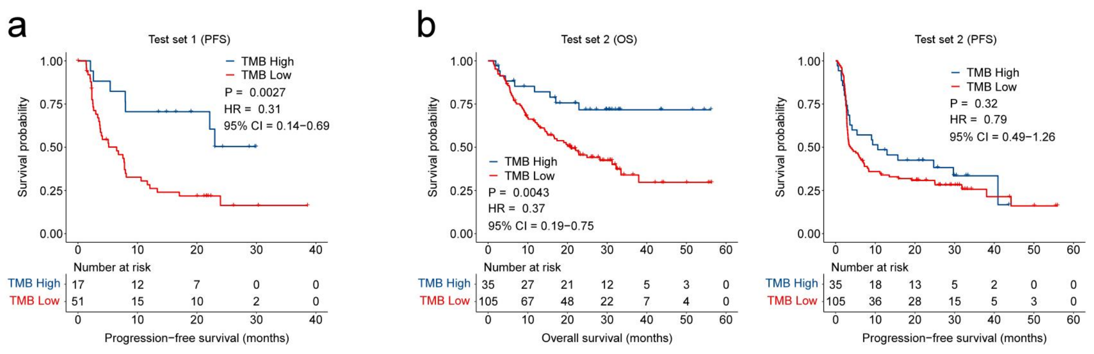


**Fig. S1. KM curves of the test sets based on the TMB (25%) cutoff groupings. a**, KM curve of PFS on test set 1 (Hellmann cohort). **b,** KM curves of OS and PFS on test set 2 (Liu cohort).

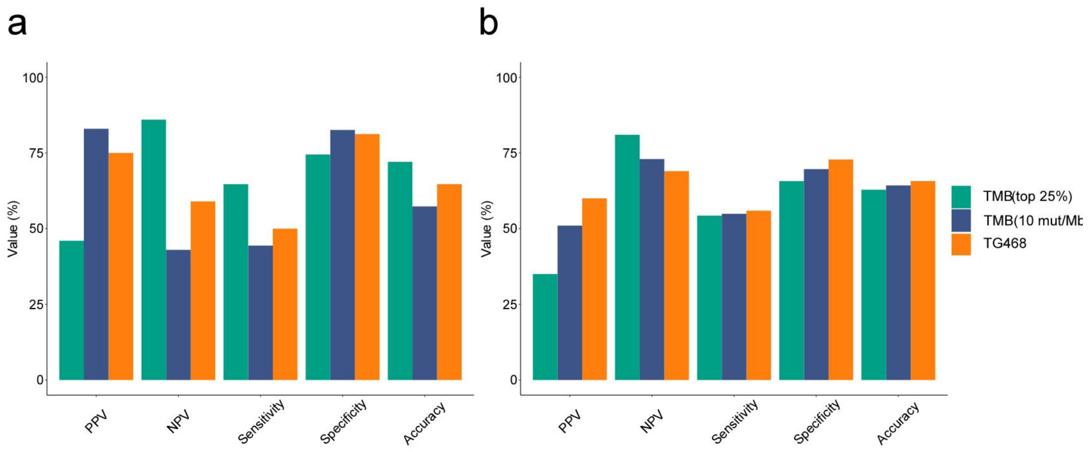


**Fig. S2. Comparisons between TG468 and TMB were made on PPV, NPV, sensitivity, specificity, accuracy. a,** Comparison of prediction performance between TG468 and TMB on the test set of Hellmann cohort. **b,** Comparison of prediction performance between TG468 and TMB on the test set of Liu cohort. Positive predictive value, PPV; Negative predictive value, NPV.

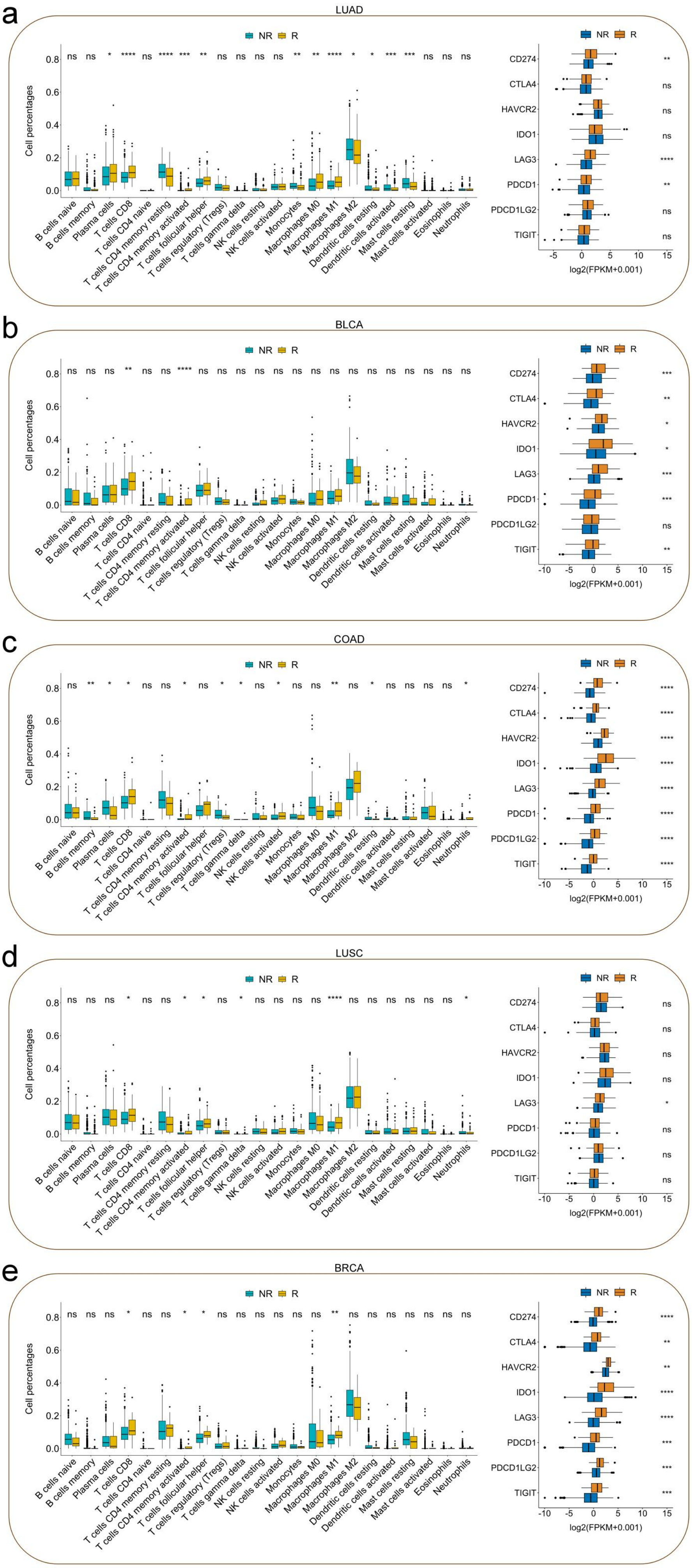
a
LUAD
NR R
ns ns * **** ns **** *** ** ns ns ns ns ** ** **** * * *** *** ns ns ns
Cell percentages
0.0 0.2 0.4 0.6 0.8
B cells naive
B cells memory
Plasma cells
T cells CD8
T cells CD4 naive
T cells CD4 memory resting
T cells CD4 memory activated
T cells follicular helper
T cells regulatory (Tregs)
T cells gamma delta
NK cells resting
NK cells activated
Monocytes
Macrophages M0
Macrophages M1
Macrophages M2
Dendritic cells resting
Dendritic cells activated
Mast cells resting
Mast cells activated
Eosinophils
Neutrophils
CD274 **
CTLA4 ns
HAVCR2 ns
IDO1 ns
LAG3 ****
PDCD1 **
PDCD1LG2 ns
TIGIT ns
-5 0 5 10 15
log2(FPKM+0.001)
b
BLCA
NR R
ns ns ns ** ns ns **** ns ns ns ns ns ns ns ns ns ns ns ns ns ns ns
CD274 ***
CTLA4 **
HAVCR2 *
IDO1 *
LAG3 ***
PDCD1 ***
PDCD1LG2 ns
TIGIT **
-10 -5 0 5 10 15
log2(FPKM+0.001)
c
COAD
NR R
ns ** * * ns ns * ns * * ns * ns ns ** ns * ns ns ns ns *
CD274 ****
CTLA4 ****
HAVCR2 ****
IDO1 ****
LAG3 ****
PDCD1 ****
PDCD1LG2 ****
TIGIT ****
d
LUSC
NR R
ns ns ns * ns ns * * ns * ns ns ns ns **** ns ns ns ns ns ns *
CD274 ns
CTLA4 ns
HAVCR2 ns
IDO1 ns
LAG3 *
PDCD1 ns
PDCD1LG2 ns
TIGIT ns
e
BRCA
NR R
ns ns ns * ns ns * * ns ns ns ns ns ns ** ns ns ns ns ns ns ns
CD274 ****
CTLA4 **
HAVCR2 **
IDO1 ****
LAG3 ****
PDCD1 ***
PDCD1LG2 ***
TIGIT ***

**Fig. S3. Analysis of TG468's prediction results on pan-cancer datasets. a-e**, Immune infiltration analysis and immune checkpoint-related gene expression analysis based on prediction results of pan-cancer datasets. Abbreviations: LUAD, Lung adenocarcinoma; BLCA, Bladder Urothelial Carcinoma; COAD, Colon adenocarcinoma; LUSC, Lung squamous cell carcinoma; BRCA, Breast invasive carcinoma.
